# Flying Without a Net: Space Radiation Cancer Risk Predictions without a Gamma-Ray Basis

**DOI:** 10.1101/2021.10.10.21264796

**Authors:** Francis A. Cucinotta

## Abstract

It is well known that the spatial distribution of ionization in cells and tissue from heavy ions and other high linear energy transfer (LET) radiation leads to qualitative and quantitative differences in biological effects compared to low LET radiation such as gamma-rays. However, models used to estimate risks involve extensive use of gamma-ray data, including low LET radiation epidemiology, the role of gamma-rays in estimates of quality factors (QF), and the dose and dose-rate reduction effectiveness factor (DDREF). In tumor induction studies, high LET radiation typically have demonstrable dose responses in many animal strains and tissue, while gamma-ray exposures often lead to a weak or poorly determined dose response at low to moderate doses (<2 Gy) leading to large uncertainties in QF estimates. Here we consider an alternate risk prediction approach, avoiding low epidemiology, the QF and DDREF, by formulating a fluence based track structure model of excess relative risk (ERR) with parameters estimated from animal studies with heavy ions and neutrons for the induction for lung and breast cancer in females and liver cancer in males. The ERR model is applied directly with cancer rates for the US population to predict lifetime risks to astronauts at solar minimum. Results for male liver and female breast cancer risk show that the ERR model agrees fairly well with estimates of a QF model with estimates of non-targeted effects (NTE), and is about 2-fold higher than the QF model that ignores NTE. The effective damage area derived by the ERR model for breast and liver tumors is several times that of a mammalian cell nucleus, which suggests NTE likely contribute to cancer risk. For female lung cancer risk, the ERR model predicts 2-fold and 5-fold lower risk compared to the QF models with or without NTE, respectively. We suggest that the direct ERR approach when coupled with improved experimental models of tissue specific cancers representing human risks would lead to large reductions in the uncertainties in space radiation risk projections by avoiding low LET uncertainties.

## INTRODUCTION

Space travel was preceded by human exposures to X-rays and gamma-rays, including medical patients, the atomic bomb detonations in Hiroshima and Nagasaki, Japan, and the use of nuclear energy. Findings from human epidemiology studies of low LET radiation (gamma-rays or X-rays) most frequently the Lifespan Study (LSS) of atomic-bomb survivors (BEIR 2006) have been the basis of predictions of cancer risks from space radiation (NCRP 1989, NCRP 2000), which are applied using a quality factor (QF) to weight organ doses for the purpose of extrapolating low LET epidemiology data to space radiation exposures. Age specific hazard rates are then formulated using background cancer rates in the population of interest, with this approach denoted as the conventional risk estimation model. Radiation epidemiology data has been invaluable in identifying the most radiogenic cancers (leukemia, breast, lung, etc.) and differences in risks with age at exposure and sex. However, the use of epidemiology data in this manner leads to a variety of uncertainties including statistical, dosimetry errors, and differences in the population makeup and time periods compared to the exposure group of interest, as well as radiation quality and dose-rate effects. A large part of the uncertainty in QFs is due to the variability in gamma-ray responses for tumor induction or surrogate cancer endpoints (Cucinotta 2015). Experimental studies of cancer induction by gamma-rays rays in mouse and rat strains are often inconsistent with some strains showing a weak or non-demonstrable dose response (NCRP 1990), which limits the strains and tissues that are available to estimate QFs. In contrast, high LET radiation has been shown to lead to demonstrable dose responses in a large number of mouse and rat strains (Ullrich 1984, Storer and Fry 1995, Di Majo et al. 1994, NCRP 1990). Also, the qualitative differences in biological effects between high and low LET radiation are not considered in QF based models.

The conventional model to cancer risk assessment is based on a hazard rate parameterized to epidemiology data, which is scaled to the background cancer rates in the population of interest, and the QF and a DDREF estimate from gamma-rays. This leads to a model for the hazard rate with several parameters to be estimated for each tissue (T) at risk based that is applied to predict proton and heavy ion cancer risk (incidence (I) or mortality (M)) (Cucinotta 2015, Cucinotta et al., 2017):

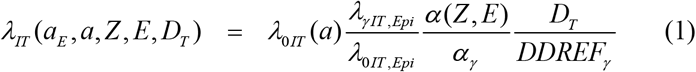

where *a* is age, *a*_*E*_ age at exposure, *Z* and *E* the ion charge number and kinetic energy, *D*_*T*_, the tissue absorbed dose. The rates, *λ*, on the right and side of equation (1) are the background age and tissue specific cancer rate in the population where risks are to be estimated, and the background and gamma-ray cancer induction rates estimated from the population used in epidemiology studies. The middle quotient in Eq. (1) is the relative biological effectiveness (RBE), which is the ratio of estimates of linear dose response coefficients for the ions and gamma-rays, or alternatively the QF. The denominator of the last quotient is the dose and dose-rate reduction effectiveness factor (DDREF) for gamma-rays. The DDREF is estimated as a ratio of acute to low dose-rate exposure effects, or from the amount of curvature in the acute gamma-ray response for cancer induction (BEIR 2006, Hoel 2015, Cucinotta et al., 2017).

The main purpose of the present report is to consider an alternate model independent of gamma-ray data that could lead to accurate space radiation risk predictions. The approach is based on direct application of the excess relative risk (ERR) from experimental studies and is denoted as the DERR model. In the DERR model the hazard rate is estimated as:

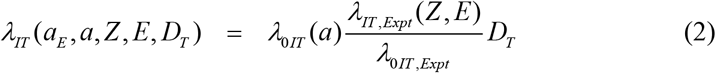

where the ratio in Eq. (2) is the ERR. The apparent advantage of Eq. (2) over Eq. (1) is the removal of uncertainties in gamma-ray effects for space radiation predictions, however as such relies on experimental findings with heavy ions and other high LET radiation in a distinct manner from the conventional model. The model is de-coupled from radiation epidemiology data although observations from these studies can be included in a distinct manner than the conventional approach as described below. Equation (2) has to consider age at exposure or time since exposure (latency) in a distinct manner than the conventional model of Eq. (1) due to this decoupling from epidemiology data. However, for adults of age 30 to 55 y typical of the ages of astronauts and for estimation of life-time risks it is not likely a large hurdle in the DERR approach. A large part of the age dependence above age 30 y is due to the life-table (declining years remaining with increasing age of exposure). In addition, inferences from radiation epidemiology data can be used to estimate the effects of age at exposure.

**Table 1** compares sources of uncertainty in the conventional model to the DERR approach. Dosimetry errors have been well studied in the LSS leading to errors >15% (BEIR VII), while retrospective organ dose assessments in other exposed populations often have larger errors. Dose estimation at particle accelerators carries small measurement errors (<5%). Other sources of uncertainty include the scarcity of ion studies on tumor induction including number of ions considered and need for studies at low doses (<0.1 Gy). However, these sources of uncertainty are similar in both approaches, and is largely an investment issue that could be overcome with larger experimental efforts than in the past. Space dosimetry, which involves prediction of organ doses and the particle charge and energy spectra contributing to doses, is estimated to have modest uncertainties (<15%) that impact both the conventional and DERR models. An important challenge for any risk prediction model is to understand if experimental models are accurately reflecting radiation cancer risks in humans. So, this is a challenge that occurs in relying on the use of QFs as well as the DERR model to estimate human radiation cancer risk.

**Table 1.** Comparisons of sources of uncertainty in conventional model to DERR model.

| **Source of Uncertainty** | **Conventional Model** | **DERR Model** |
| --- | --- | --- |
| **Statistical** | Epidemiology data, experimental for gamma-rays, experimental for ions. | Experimental for ions alone. |
| **Dosimetry** | Epidemiology data, and in accelerator studies with animals. | In accelerator studies with animals. |
| **Time period** | Epidemiology data | None |
| **Genetic and environmental factors** | Likely distinct in epidemiology data from modern day radiation workers including astronauts.<br>Considered in experimental design for gamma-rays and ions. | Considered in experimental design for ions. |
| **Dose-rate** | Extrapolation of epidemiology, data, and experimental data for gamma-rays to risks of ions to chronic low doses. | Extrapolation of experimental data for ions to chronic low doses. |
| **Range of dose and ion types considered** | Limited by investments in experiments performed. | Limited by investments in experiments performed. |
| **Extrapolation of tissue specific factors from mouse to humans** | For both gamma-rays and ions. | For ions. |

The NASA Space Cancer Risk (NSCR) model (Cucinotta 2015, Cucinotta et al. 2017, 2020) is an extension of the conventional approach to include QF’s based on microscopic energy deposition and ion track structure concepts, while making a detailed uncertainty analysis of the various components that enter into risk assessments to estimate an overall uncertainty. In this report we consider results of tumor studies in mice and rats from heavy ions and fission neutrons to develop a fluence dependent model of ERR for breast, lung (female) and liver (male) cancer risk. Fission neutrons are a high LET form of radiation with the majority of its effect due to high LET protons with energies below 1 MeV. The model considers published studies in several mouse strains for a small number of heavy ion types or fission neutrons, and uses the inferences from the NSCR radiation quality model to predict breast, lung and liver cancer risks for other ion types and energies. High LET radiation dose responses are observed to saturate at higher doses leading to a bending of dose response curves at doses of about 0.2 to 0.6 Gy dependent on tumor type. The saturation is likely due to a combination of cell kill, repopulation, and stress and immune responses (Ullrich 1983). We consider both a linear in dose ERR model and an ERR model with a bending dose response leading to saturation at high dose. This model reduces to a linear response at very low dose. The models are cast in terms of charged particle fluence and compared to experiments and used to make predictions for galactic cosmic ray (GCR) exposures.

## METHODS

We fitted data for the dose response for heavy ion or fission neutron induced lung, mammary and hepatocellular tumors in mice or rats using a linear function or the function

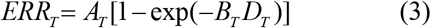

This function describes the bending over and saturation of high LET dose responses observed in experiments. The parameter *A*_*T*_ is seen to represent the saturation value of the ERR, while at sufficiently low dose the ERR becomes linear in dose with coefficient, *A*_*T*_ *x B*_*T*_.

Equation (3) is then recast as the ERR in terms of tissue specific ion fluence, *F*_*T*_, for charge number, Z and kinetic energy, Z as:

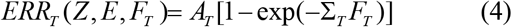

The normalization parameter, Σ_0T_ (in units of area, μm^2^) of the effective action cross section is determined by B_T_ and considering the details of the energy spectra in the mouse tumor induction experiments with neutrons or heavy ions. The effective action cross section, Σ_T_, in Eq. (4) is represented by the form:

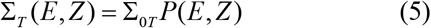

We use a mathematical form motivated by track structure models in the NSCR model:

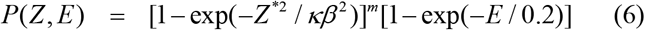

where *m* and *κ* are model parameters, β is the ion velocity scaled to the speed of light, and Z* is the effective charge number that describes electron pickup as the ion’s velocity is decreased. The function *P(E,Z)* allows us to extrapolate from ions considered in experiments to other ions of a given Z and E based on a large number of prior studies in mice and cell culture models (Cacao et al. 2016; Cucinotta at al. 2017) to estimate values of the parameters κ and *m*.

We use the above ERR descriptions to predict the risk of exposure induced cancer (REIC) for annual GCR exposures, and performed uncertainty analysis using Monte-Carlo methods as described previously (Cucinotta et al., 2013; Cucinotta et al., 2017).. The λ_I_ (or λ_M_) is a sum over rates for each tissue that contributes to cancer risk, λ_IT_ (or λ_MT_). The REIC is calculated by folding the instantaneous radiation cancer incidence-rate with the probability of surviving to time *t*, which is given by the survival function *S*_*0*_*(t)* for the background population times the probability for radiation cancer death at previous time, summing over a space mission exposure, and then integrating over the remainder of a lifetime.

Lifetables from the U.S. Center of Disease Control and Prevention (CDC) for male and females in the US are used in calculations (CDC, 2020). Tissue specific cancer incidence and mortality rates are available from National Cancer Institute, Surveillance Epidemiology and End Results (SEER) program with the latest data collected for the years 2014-2018, and provide race and ethnic group, age, and sex specific rates. For cancer incidence we used the SEER delay-adjusted rates, which estimate the impact of delay in reporting of cancer cases (Midthune et al., 2005, SEER 2021).

### Fission Neutron and Space Radiation Organ Exposures

To apply the models described above the charged particle energy spectra are needed. Laboratory based heavy ion experiments are assumed to be in the track segment model neglecting nuclear fragmentation in the beam line (Pak and Cucinotta, 2021). Fission neutrons have energies largely below 5 MeV which simplifies calculation of particle spectra to the elastic recoils produced in tissue (H, C, O) and gamma-rays. Based on earlier findings of Storer et al. (1979), the experiments of Ullrich (1984) and Storer and Fry (1995) ignore gamma-ray contributions to absorbed doses due to the much larger biological effectiveness of neutrons with energy < 5 MeV. We followed this approach for the various experiments considered using neutrons. The charged particle spectra for a ^252^Cf fission source developed by Dennis and Edwards (1975) are used for calculations.

GCR exposures include primary and secondary H, He and HZE particles, and secondary neutrons, mesons, electrons, and γ-rays over a wide energy range. We used the HZE particle transport computer code (HZETRN) with quantum fragmentation model nuclear interaction cross sections (Wilson et al. 1994; Cucinotta et al., 2007; Kim et al. 2015) to estimate particle energy spectra for particle type *j, φ*_*j*_*(Z,E)* as described previously Cucinotta et al., 2017). GCR organ dose equivalent show little variation from 10 to 50 g/cm^2^ of shielding, and we use 20 g/cm^2^ for calculations, which is a typical average shielding amount.

A mixed-field action cross section is formed by weighting the particle flux spectra, *φ*_*j*_*(E*) for particle species, *j*, contributing to GCR exposure evaluated with the HZETRN code with the pseudo-biological action cross section for mono-energetic particles and summing over all particles and kinetic energies:

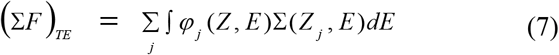

## RESULTS

We fit the model of Equation (3) and a linear dose response model for the ERR for several data sets in mice or rats for hepatocellular, lung and mammary carcinoma. We chose data sets (**Table 2)** with sufficient data with more than one animal strain for high LET radiation at doses <0.5 Gy that considered lifespan studies or with sufficient follow-up time that represents most of the animal’s lifespan. This approach avoids a parametrization influenced by reduced latency at early time points for high LET exposures. Because of restricted follow-up time and slower progression of spontaneous tumors compared to radiation induced tumors, ERR values will have a strong age dependence for limited follow-up times (Cucinotta and Wilson, 1995). Of note is the Harderian gland prevalence studies ERR >10 occur for Fe particle at 600 d prevalence. However, the spontaneous tumor rate increases from ∼2.9% at 600 days to ∼10 percent at 800 days (Fry 1993, Cucinotta and Wilson, 1995), and a declining ERR with time after exposure occur suggesting age the use of an age adjusted ERR (Ullrich et al., 1977). A similar observation is well known in experiments with Sprague-Dawley (SD) rats for induction of mammary tumors (benign or malignant) where a sufficient time shift to earlier appearance for high LET radiation occurs (NCRP 1990).

**Table 2.** Sources of tumor data in mice and rats exposed to fission neutrons (FN) or heave ions. Data is chosen as representative of major cancer types in humans where more than one animal strain was reported at doses of high LET radiation below 0.5 Gy, and with observation period for tumor appearance representative of animal lifespan.

| **Model** | **Tumor** | **Age at irradiation** | **Irradiation Types** | **Duration of observations** | **Reference** |
| --- | --- | --- | --- | --- | --- |
| BALB/c/AnNBdf Female mice | Mammary adenocarcinoma | 120 d | FN mean energy 2 MeV, acute and chronic | Lifespan | Ullrich 1984 |
| BALB/c/AnNBdf Female mice | Lung adenocarcinoma | 120 d | FN mean energy 2 MeV, acute and chronic | Lifespan | Ullrich 1984 |
| CBA/CaJ Male mice | Hepatocellular carcinoma | 8-14 weeks | $^{56}\text{Fe}$ (1 GeV/u), acute | Observed to age 800 d | Weil, et al. 2009 |
| C3H/HeNCRl Male mice | Hepatocellular carcinoma | 8-10 weeks | $^{56}\text{Fe}$ (0.6 GeV/u),<br>$^{28}\text{Si}$ (0.3 GeV/u), acute | Observed to age 800 d | Weil, et al. 2014 |
| Sprague-Dawley Female rats | Mammary carcinoma | 7 weeks | Fast neutrons mean energy 2.3 MeV, acute | Observed to age 90 weeks | Imaoka et al. 2017 |
| B6CF1 (C57BL/6 Bd x BALB/c Bd) Male mice | Hepatocellular carcinoma | 16 weeks | FN mean energy 2 MeV, acute and chronic | Observed to 1200 d | Storer and Fry, 1995 |
| B6CF1 (C57BL/6 Bd x BALB/c Bd) Female Mice | Lung Adenocarcinoma | 16 weeks | FN mean energy 2 MeV, acute and chronic | Observed to 1200 d | Storer and Fry, 1995 |
| BCF1 male mice | Hepatocellular carcinoma, all solid cancers | 3 months | FN mean energy 0.4 MeV, acute and fractionated | Lifespan | Di Majo et al. 1994 |

**Figure 1** shows results for hepatocellular carcinoma for several mouse strains. Results for heavy ions in CBA and C3H mice (**Fig. 1 panels A and B**) are similar as shown by the parameter values listed in **Table 3**. The ERR saturation model led to improved fits compared to a linear response model. Experiments in B6CF1 male mice analyzed by Storer and Fry (1995) considered low doses of acute and fractionated fission neutrons (mean energy ∼2 MeV), however because the results were not significantly different we pooled the data in our fits. Here the linear model fit better (**Fig. 1, panel C**), however the lack of higher doses in the study does not allow for a study of a bending in the dose response or for a possible saturation of response at higher doses. **Figure 1, panel D** shows the results for ions in CBA and C3H mice and for fission neutrons of lower mean neutron energy (0.45 MeV) by Di Majo et al. (1994). For these comparisons, only the neutron dose estimate is useed. **Figure 1, panel D** includes the application of the ERR model of equation (3) using Eq. (7) for Si, Fe and fission neutrons, which show very similar responses. The Fe particles are more efficient per unit fluence (per particle), however at identical doses, the fluence of recoil particles from fission neutrons or Si ions is about 2-fold higher than Fe.

**Table 3.**
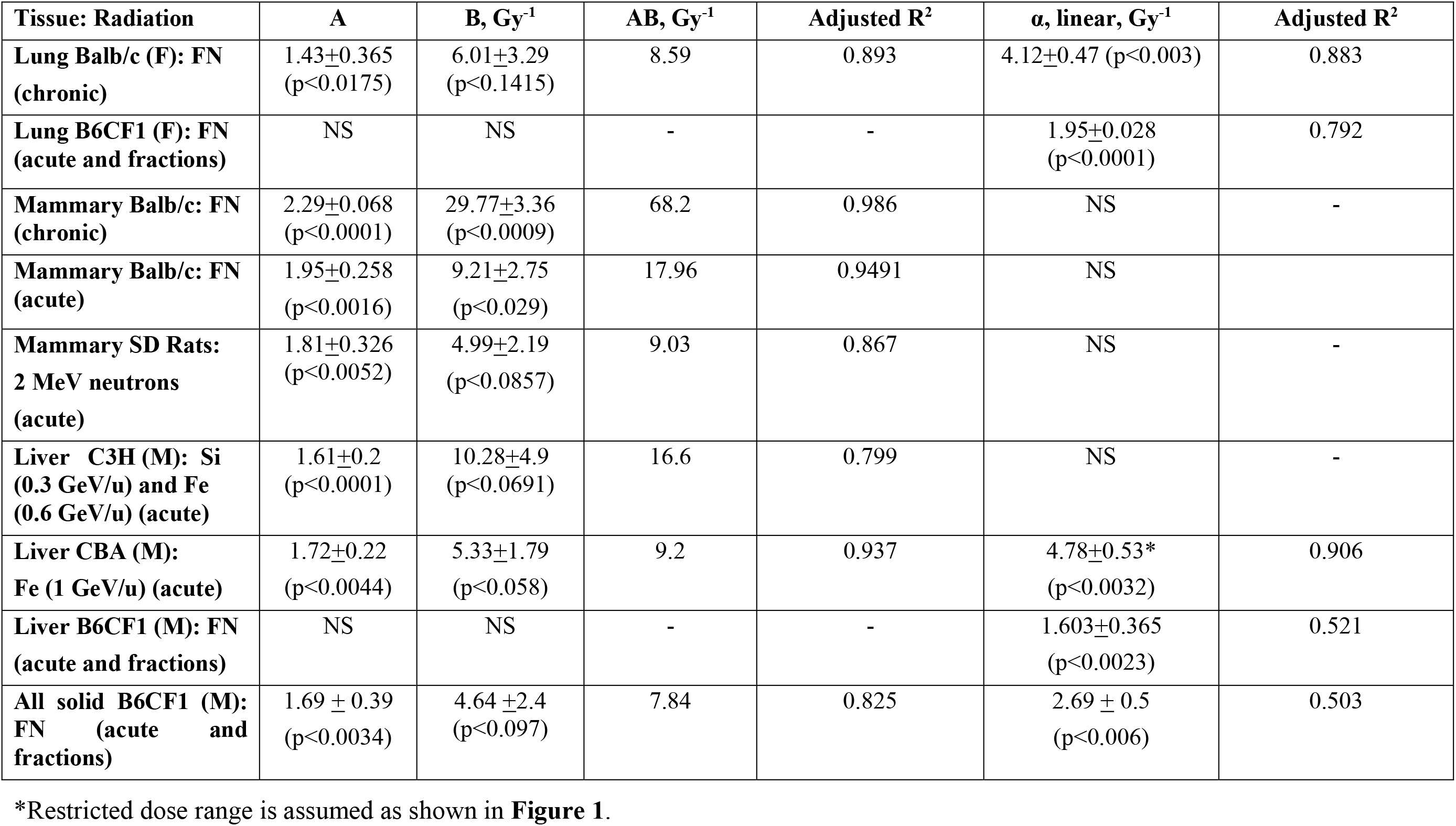
Parameter estimation for ERR function for lung and mammary tumors in female and liver tumors in males in several strains of mice or rats. Model functions are ERR = A [1-exp(-B Dose)] or ERR = α Dose. Means, standard deviations and adjusted R^2^ values are listed. Abbreviations NS=Not significant, SD = Sprague-Dawley, FN = fission neutrons.

**Figure 1.**
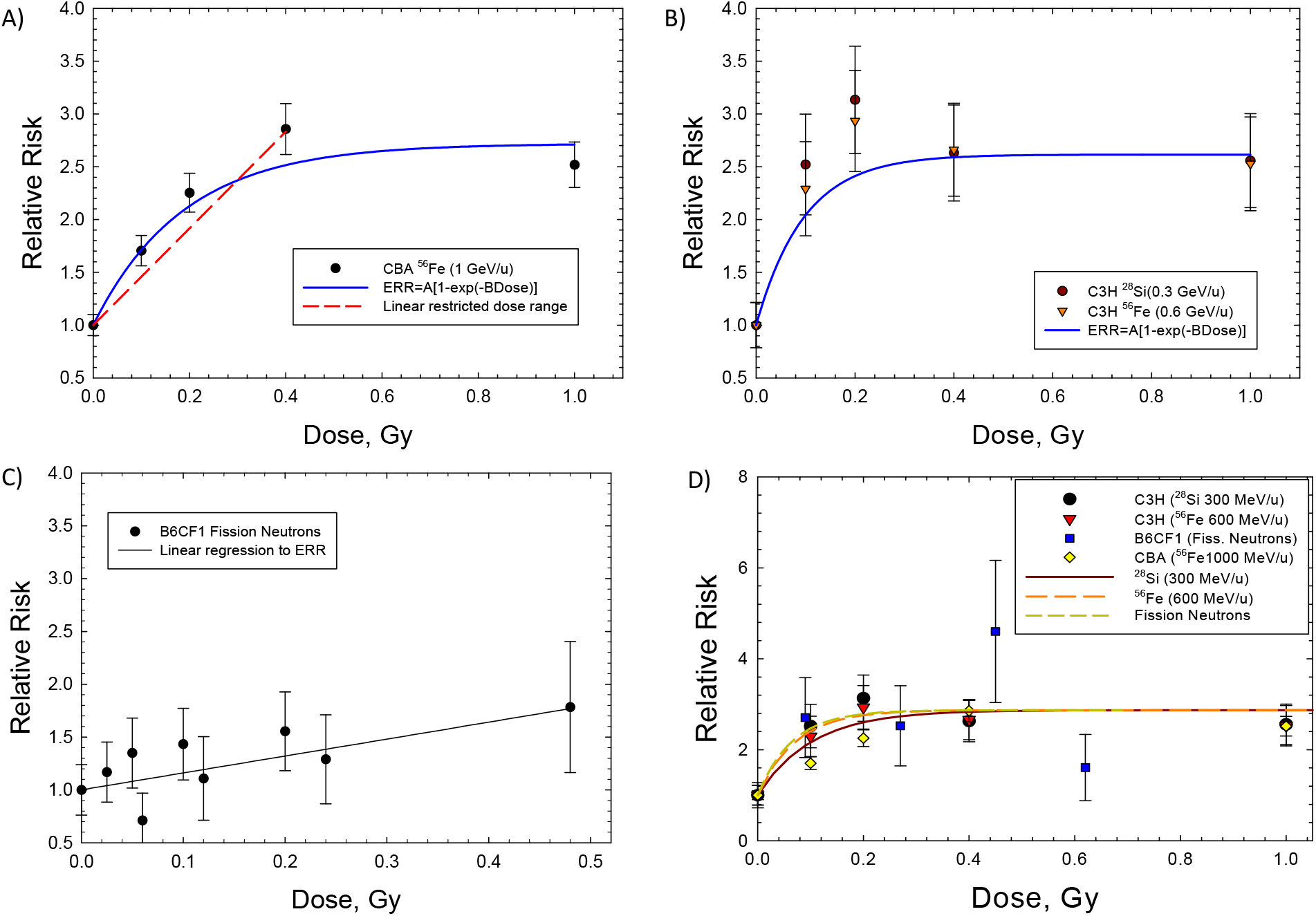
Dose response for hepatocellular carcinomas in several strains of male mice exposes to heavy ions (HIs) or fission neutrons (FN). The saturable ERR model described in the text is compared to the experimental data. A) CBA exposed to HIs. B) C3H exposed to HIs, C) B6CF1 exposed to FN. D) Model fits to several strains of mice showing similarity of HI and FN responses.

Dose response data for induction of lung adenocarcinoma were reported for female Balb/C and B6CF1 mice for acute and chronic (or fractionated) fission neutron exposures (**Figure 2 Panel A and Panel B**). The saturation ERR model provided a better fit to data for Balb/c mice, while the linear model was a better fit to the B6CF1 mice data. Figure 2 Panel C shows results in B6CF1 mice for female and male mice. For lung carcinomas in mice there is data for cyclotron neutrons in male and female SAS/4 mice (Coggle, 1988) (**Figure 2 Panel D**), however these data present the complication of inelastic scattering by neutrons and the use of thorax only irradiation. Also, cyclotron neutrons will have a lower effective LET compared to fission neutrons or Si and Fe particles which limits a direct comparison to heavy ion results. The use of partial body irradiation leads to a higher RR compared to the other experiments. For both B6CF1 and SAS/4 mice female mice have higher ERR values compared to males.

**Figure 2.**
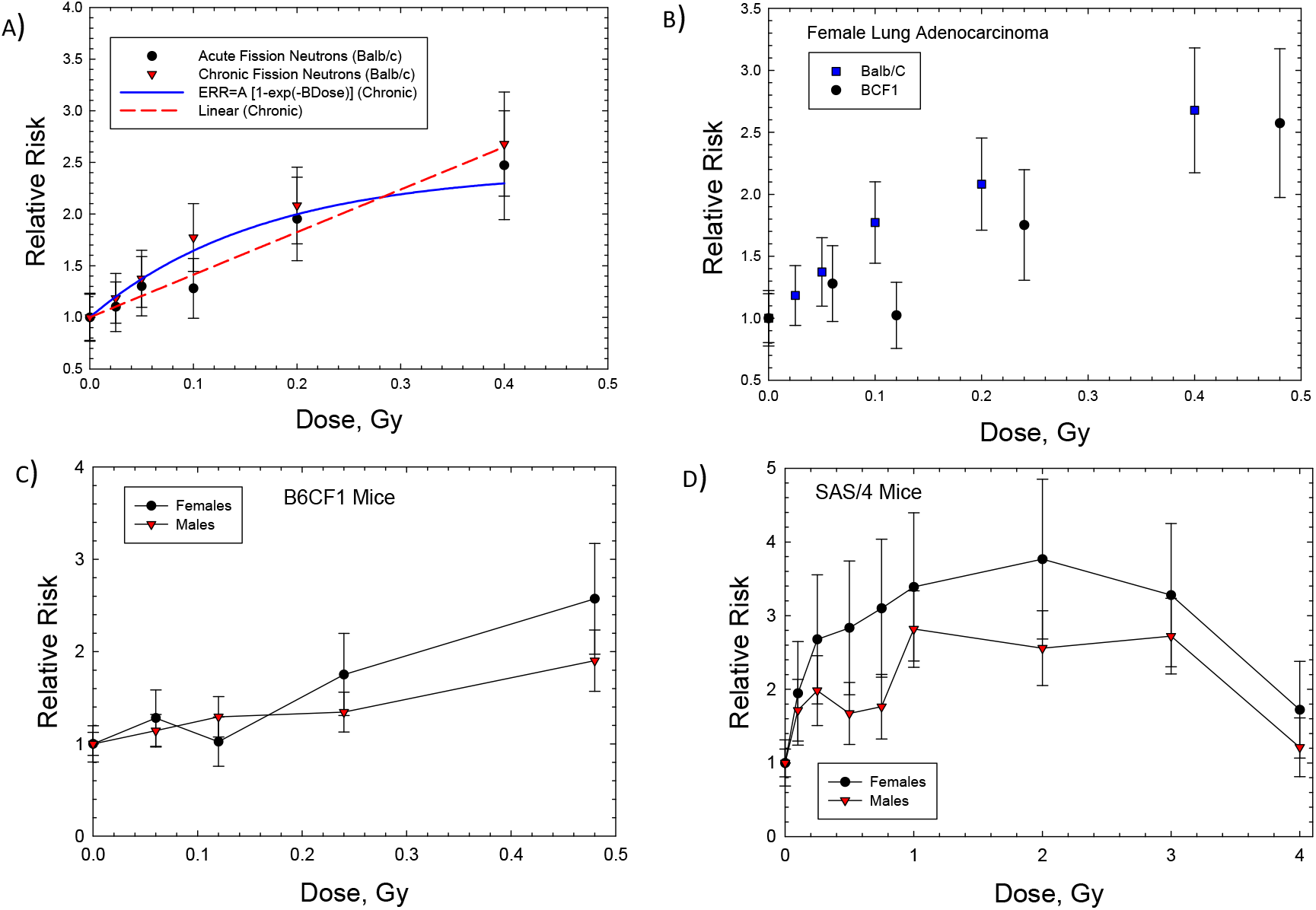
Dose response for Relative Risk of lung adenocarcinomas in female Balb/c, B6CF1, or SAS/4 mice exposed to neutrons. A) Female Balb/c mice exposed to fission neutrons (FN) showing linear and saturable ERR models. B) comparison of Balb/c to B6CF1 mice exposed to FN. C) Comparison of female to male B6CF1 mice exposed to FN. D) Comparison of female to male SAS/4 mice exposed (thorax only) to cyclotron neutrons.

For mammary tumors in mice, results for Balb/C mice are the only available dose response data for high LET radiation. Storer and Fry (1995) note that the B6CF1 strain is not susceptible to radiation induced mammary tumors. However, there are several data sets for SD rats with neutrons (Vogel 1969, NCRP 1990, Dicello 2005). We considered the recent data of Imaoka et al. using 7-week-old rats exposed to a neutron spectrum with mean energy of 2 MeV because of the long follow-up period of 70 weeks approaching a lifespan study. **Figure 3** and **Table 3** show that the saturable ERR model fits the acute data in Balb/c mice and SD mice with similar saturation values with a stronger response in Balb/c. An ERR enhancement is observed for the chronic exposures in Balb/c mice (Ullrich 1984) with a higher saturation value of 2.29 compared to 1.95 in the acute exposures with fission neutrons. Fits to the various mammary tumor data with a linear ERR model did not lead to a significant result. A similar saturation of response in SD rats is seen by Dicello et al. (2004) with Fe particle exposures reporting excess incidence for combined carcinomas and fibromas, however data on the spontaneous tumor rates were not reported to make a numerical estimate of ERR.

**Figure 3.**
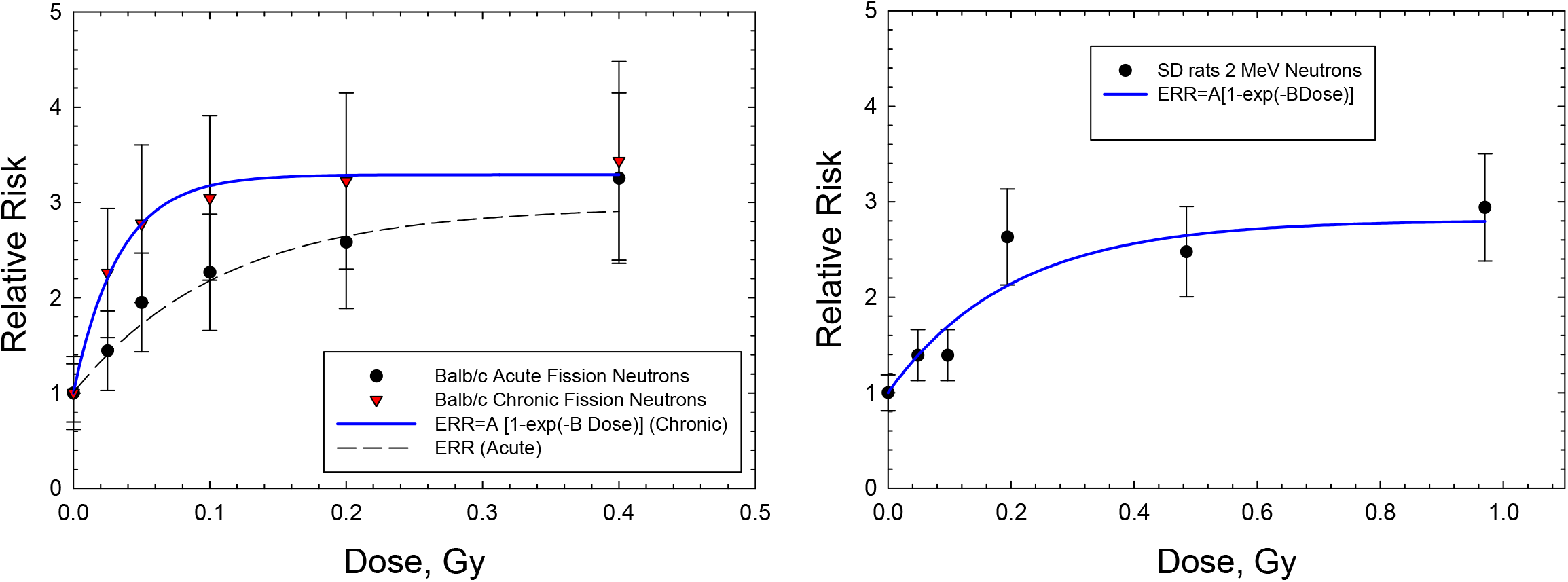
Dose response for Relative risk of mammary adenocarcinomas in Balb/c mice and Sprague-Dawley rats exposes to fission neutrons or neutron source with average energy 2 MeV. The saturable ERR model described in the text is compared to the experimental data.

Di Majo et al. (1994) also reported dose response data for all solid cancers. Dose fractionation using 5 daily doses were used in the experiments. **Figure 4** shows that the saturable ERR model provides a good fit to these data. Data for all solid cancers were not reported for B6CF1 mice by Storer and Fry (1995).

**Figure 4.**
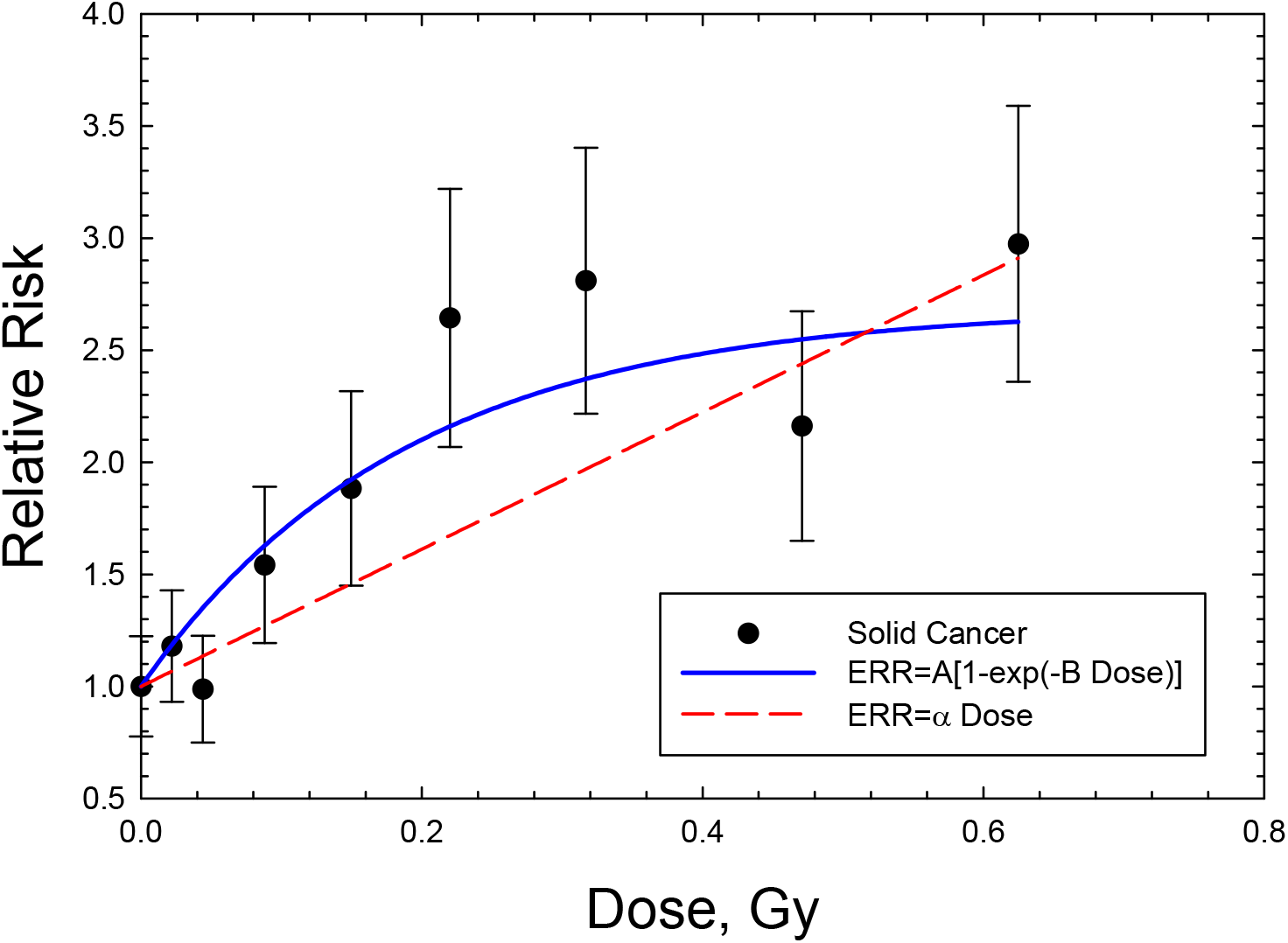
Dose response for Relative Risk of solid cancers in male B6CF1 mice to fission neutrons (mean energy 0.4 MeV). The saturable ERR model described in the text is compared to the experimental data.

We made subjective choices of parameters to the various fits in **Table 3** that allow us to make predictions of cancer risks for GCR and compare to the NSCR-2020 model. The preferred parameter values are shown in **Table 4**. For the uncertainty in the A_T_ and Σ_T_ parameters we assumed normal distributions with standard deviation of 33% of the mean. The analysis of Storer and Fry (1995) are restricted to lower doses which preclude observation of a likely saturation effect, while the initial slopes are also influence by bending ever for doses as low as 0.1 Gy of high LET radiation. Therefore, we focus on the saturation ERR model for space radiation calculations as described below. For mammary cancers the protracted exposures used by Ullrich (1984) are preferred, however parameter values were adjusted to reflect the average of the acute exposures in Balb/c mice and SD rats.

Prediction for an annual GCR exposure for missions at average solar minimum conditions are shown in **Table 4** for 35-y and 50-y old astronauts. Predictions include those for breast and liver cancer in females and liver cancers in males. The NSCR-2020 (Cucinotta et al. 2020) model is used to make predictions with and without an estimate of NTE’s on the QF. Interestingly the results for breast and liver cancers with NTEs agree closely to that of the DERR model. The age dependence is similar since the largest contributor above age 30-y to a declining risk with increasing age of exposure is the constraint of the life table, which is identical in the models presented using US population data. Predictions for lung cancer risks in females the DERR suggest about a 2-fold and 5-fold lower risk compared to NSCR-2020 with and without NTE, respectively. Predictions of cancer risks for the US population for females are higher than males for several types of cancer including stomach, liver, urinary track and all solid cancers, however the largest difference is for females with a F:M ratio of 2.83 (Brenner et al. 2018). It is not clear if this difference is reflected in mouse experiments. A higher ERR for females compared to males occurs for lung cancer in thorax only irradiation of cyclotron neutrons in SAS mice (Coggle 1998) and whole-body exposures to FNs in B6CF1 mouse (Storer and Fry, 1995). These differences will be considered in future work.

**Table 4.** DERR model preferred parameters for cancer risk in several tissues.

| **Tissue** | **A<sub>T</sub>** | **B<sub>T</sub>, Gy<sup>-1</sup> (Σ<sub>0</sub>, μm<sup>2</sup>)** |
| --- | --- | --- |
| Lung (Females) | 1.4 ± 0.4 | 5 ± 2 (250) |
| Liver (Males) | 1.65 ± 0.2 | 7.5 ± 3 (400) |
| Breast (Females) | 2.1 ± 0.3 | 20 ± 5 (1000) |

**Table 5.** Predictions of Risk of Exposure Induced Cancer (REIC) for 1-year exposures with average solar minimum conditions for astronauts of 35 and 50 y at mission launch. Results for the NSCR-2020 mode with and without non-target effects (NTE) are shown in comparison to the DERR model. Uncertainties for physics and the *κ* and *m* parameters are the same in each prediction as described previously (2017; 2020). Uncertainties in other parameters are distinct for the various predictive models.

| Cancer | NSCR-2020 Model | NSCR-2020 with NTE | NSCR-DERR Model |
| --- | --- | --- | --- |
|  | %REIC for Mission Age 35 years |  |  |
| Lung Females | 1.98 [0.52, 6.0] | 4.7 [1.4, 10.7] | 0.92 [0.2, 3.1] |
| Breast Females | 4.2 [0.81, 13.2] | 9.7 [2.2, 23.5] | 8.5 [2.1, 21.5] |
| Liver Males | 0.11 [0.03, 0.34] | 0.27 [0.08,0.63] | 0.24 [0.05, 0.61] |
|  | %REIC for Mission Age 50 years |  |  |
| Lung Females | 1.99 [0.51, 6.03] | 4.68 [1.4, 10.7] | 0.88 [0.17, 2.89] |
| Breast Females | 2.98 [0.57, 9.41] | 7.0 [1.53,17.2] | 6.8 [ 1.67, 16.2] |
| Liver Males | 0.07 [0.02, 0.22] | 0.17 [0.05, 0.40] | 0.2 [0.05, 0.44] |

## DISCUSSION

Approaches to extrapolate from experimental data to humans have been limited with the conventional approach using QFs the mainstay of space radiation risk predictions for several decades (NCRP 1989, NCRP 2000, Cucinotta et al. 2013). The NCRP considered extrapolation approaches for space radiation in the past, including fluence based risk coefficients and microdosimetric approaches (NCRP 2001). However, these approaches relied on gamma-ray data for predictions. Relative risk models were considered in a NCRP Report (NCRP 2005) on extrapolation from nonhuman to humans. Carnes developed an approach to extrapolate mortality after radiation exposure between animal models and humans focusing on age specific survival with a linear dose response model (Carnes et al., 1998, NCRP 2005). A Cox proportional hazard model describing underlying intrinsic and extrinsic causes of mortality was successful in predicting risks in several species. However, with no consideration of specific aspects of tissue specific risks, ion track structure or deviation from a linear response for high LET radiation.

In this paper we considered direct application of ERR estimates high LET tumor induction studies to risk predations independent of gamma-ray data in totality. Therefore, possible limitations in risk estimates that use gamma-rays as a basis, such as qualitative differences between high and low LET, are avoided in entirety. Because of the scarcity of heavy ion tumor dose response data, the variation in responses with ion Z and E are assumed to follow the same variation as observed in other radiobiology experiments (Cacao et al., 2016; Cucinotta et al., 2017), however calibrated to tumor induction studies through the A_T_ and Σ_T_ parameters. The mathematical forms are motivated by amorphous track structure approaches (Cucinotta et al, 1999), however with similar radiation quality dependences found using frequency distributions in nanoscale volumes (<25 nm) based on Monte-Carlo track structure simulations (Goodhead 1990, Cucinotta et al. 2013). Experiments with Z and E were the probability function of equation (6) are such that *P(Z,E)∼*0.5 will have the maximum effectiveness per unit dose, and this condition approximately holds for fission neutrons and the energies of Fe and Si ions considered. Of note is the value of Σ_T_ for breast and liver cancers, 1000 μm^2^ and 400 μm^2^, respectively are much larger than the cross-sectional area of a cell nucleus, which is suggestive of a role for NTEs (Kadhim et al., 2013; Illa-Bochaca et al., 2014).

The uncertainties in physics estimates are a minor contribution to the overall uncertainty. The major contributor to the uncertainty estimates in the DERR model is the κ and Σ_T_ parameters. This is similar to the conventional form of the NSCR model with κ and a parameter that normalizes and effective cross section the main contributors, however with the additional uncertainties in the low LET data inputs to the model. The uncertainty in the values of the κ and Σ_T_ parameter could be substantially reduced with prospective experimental design rather than the conservative estimates made here based on the scarcity of historical data.

A main focus of this report is that experimental models that are designed to accurately represent tissue specific cancer risks should play the major role in reducing uncertainties in risk projection, and also are needed for realistic countermeasure studies. The parameter values obtained suggest heterogeneity of responses in the small number of rodent models considered. Prior reviews have suggested a number of design criteria, which includes focus on the tissues which represent the largest contributors to risk, which are lung, breast, colon, stomach, liver and leukemias for adults (>30 y). In addition, considerations of cell of origin for cancer, and genetic diversity similar to humans should be considered. Radiation cancer susceptibility studied in the mouse has been shown to reflect spontaneous rates of cancer development (Fry 1981, Storer et al., 1988). A practical limitation of the ERR approach would be application to models, such as transgenic mice, that have small or negligible spontaneous cancers. However, such models are not likely to represent human risk albeit mechanistic hypothesis are effectively investigated with such approaches (Suman et al., 2016). Genetically diverse cohorts (Edmonson et al. 2020) are a possible approach however the number of dose groups (∼5 to 6) needed to accurately define a dose response for several ions is likely prohibitive due to the large number of mice used in these studies.

The main conclusion of this reports is that space radiation cancer risk predictions independent of the low LET epidemiology such as the epidemiology of the LSS, QF’s and DDREF are readily made for several tissues with existing high LET tumor data, and the results are similar to the NTE estimates of the NSCR model with the exception of lung cancer in females. In future work we will extend our analysis to data from higher energy neutrons and consider time dependent models of ERR as function of radiation quality using more detailed models of neutron transport than considered in the present report. The use of time after exposure dependent ERR models will allow for parametrizations of experiments limited by follow-up time, while the present report considered only lifespan studies. In addition, an approach to include information from low LET studies in the action cross section model will be considered.

## Data Availability

All data produced in the present work are contained in the manuscript.

## ACKNOWLEDGEMENT

The author has not conflict of interest. Funding is through the University of Nevada Las Vegas

